# Mathematical Analysis of an Age- and Socioeconomic-Stratified COVID-19 Transmission Model

**DOI:** 10.1101/2024.10.15.24315447

**Authors:** Gbeminiyi J. Oyedele, Oluwarimi J. Idowu, Michael J. Tildesley

## Abstract

The COVID-19 pandemic has sparked significant interest in developing mathematical models that capture more of the complexities of the dynamics of disease transmission and control. In this study, we presented a deterministic compartmental model for the transmission of COVID-19. The model has eight compartments: susceptible, exposed, asymptomatic, unreported symptomatic, reported symptomatic, hospitalised, recovered, and dead. Individuals in each compartment are discretised into age and deprivation deciles to study the combined effect of both factors on disease dynamics. We analyse the model and present the results for both the disease-free and endemic equilibrium states. We evaluate the basic reproduction number using a next-generation matrix approach. We also prove the local and global stability of the disease-free and endemic equilibria. Sensitivity indices are calculated both analytically and numerically to identify the parameters that have the greatest influence on *R*_0_. Our results suggest that transmission, recovery, treatment, and testing rates need to be closely monitored to reduce the disease burden. Specifically, prompt testing, treatment, and recovery are critical for reducing *R*_0_.

## 1 Introduction

COVID-19 has resulted in an unprecedented global health crisis, significant changes in the world’s social structure, and widespread economic consequences. The transmission of SARS-CoV-2 occurs mainly via respiratory droplets and direct contact with infected surfaces [1]. The disease presents with diverse symptoms, ranging from a mild cough, fever, and fatigue to severe pneumonia [1]. Understanding the dynamics of COVID-19 transmission can provide valuable information regarding the most effective intervention strategies to mitigate the ongoing impact of the virus. The aim of this study is to develop and analyse a novel mathematical model that integrates an age and deprivation-decile contact mixing structure and time-dependent testing capacity to better understand disparities in infectious disease transmission and control. By incorporating these factors into the contact structure of a compartmental model, we can evaluate their impact on disease spread, analytically study stability conditions, and identify key parameters influencing transmission dynamics. The objective is to provide actionable insights for targeted public health interventions, especially in mitigating the disparities in infectious burdens across deprivation deciles.

The transmission and control of COVID-19 have been extensively studied using mathematical models since the SARS-CoV-2 virus emerged in late 2019 [2–4].

Epidemiologists have worked closely with policymakers using various modelling techniques to provide guidance on disease outbreaks. The resulting models have been helpful in addressing several critical questions, including the projected final size of a given outbreak, the estimated number of individuals who will be infected, the timing of the peak of infection, and the optimal method for controlling the spread of the disease [5–8]. The COVID-19 outbreak has led to significant interest in developing robust mathematical models that can capture more complexities of the dynamics of disease transmission and control. Infectious disease models have proven to be essential tools in epidemiological research.

The findings of Daniel Bernoulli in the 18th century laid a strong foundation for the mathematical modelling of infectious diseases [9], and this research encouraged policymakers to adopt universal inoculation [10]. Modern mathematical modelling began in the 20th century [11–13]. In his research on malaria eradication, Ross [13] formulated a model to examine the threshold at which the mosquito population must be reduced to eliminate malaria. However, the work of Kermack [11, 12] became the foundation of modern mathematical modelling of infectious diseases with the formulation of the susceptible–infected–recovered (SIR) model. This model is useful in describing patterns in disease spread but can fail to capture the fine details required for some diseases. Over the years, this model has been adapted to incorporate more realistic scenarios that can more accurately capture the dynamics of infectious diseases.

Age structures have been widely used in infectious disease models to capture differences in disease susceptibility and contact patterns between age groups [14–17]. Kumar and Abbas [15] formulated an age-structured model to analyse infection transmission through direct contact (age-dependent mixing) and indirect contact (e.g. contaminated surfaces) using McKendrick–von Foerster partial differential equations (PDEs). Their model was formulated as an abstract semi-linear Cauchy problem, which allowed the authors to prove the existence of solutions and steady states. Chhetri *et al*. [18] used an age-structured model to study the COVID-19 transmission, vaccination, and control strategies. They analysed the model’s positivity, boundedness, stability, and basic reproduction number *R*_0_ using the next-generation matrix approach. Their numerical analysis considered various factors, including the effect of vaccination coverage and its efficacy in minimising infections and deaths. The age-structured PDE model proposed by Jing *et al*. [19] was used to study disease dynamics by incorporating vaccination and antiviral treatment as control measures. Their theoretical analysis proved the existence and stability of equilibria, showing that *R*_0_ *<* 1 leads to eradication and *R*_0_ *>* 1 leads to endemicity. However, substantial evidence indicates age-related disparities in susceptibility, severity, and mortality during the COVID-19 pandemic [16, 20, 21]. Davies *et al*. [16] found that individuals under 20 years of age are less susceptible, whereas O’Driscoll *et al*. [20] observed lower infection fatalities in the

5–9 age group than in the older groups. The age-structured model by Hilton and

Keeling [22] estimates a country-specific basic reproduction number (*R*_0_) and found that age and contact structures are crucial in COVID-19 transmission and that countries with older populations experienced a faster spread of the disease. The COVID-19 pandemic did not affect individuals and communities equally because individuals living in overcrowded conditions [23] and high-contact service occupations [24] were likely to be at a higher risk of infection than those who could work from home or live in a smaller family setting. This may increase the burden of COVID-19 in socioeconomically disadvantaged communities. Therefore, a mathematical model is essential to examine the factors influencing disproportionate disease outcomes across deprivation deciles during the COVID-19 pandemic.

Several studies have attempted to understand the impact of confounding factors on infection outcomes [25–29]. For example, Hilton and Keeling [25] considered the combined effect of age and household structure on disease outcomes; Manna *et al*. [26, 27] *developed a socioeconomic mixing matrix that combines age and other socioeconomic factors, such as income, employment, and level of education, to understand disease outcomes; and Hale et al*. [29] used a stochastic compartmental infectious disease model that incorporates age and deprivation deciles using the Bayesian approach. The study by Munday *et al*. [30] was important for the development of the combined age and deprivation-decile mixing structure used in this study. Their model combines age and two risk groups (high and low) to understand the impact of vaccination on infectious disease (influenza and rubella) burdens. Although risk groups in disease modelling are useful, they are a “wide net” (meaning that different infections have different risk groups) when it comes to stratifying the population to understand infection outcomes.

This study presents a deterministic model that incorporates the confounding effects of age and deprivation-decile contact mixing, coupled with the time-dependent testing capacity of COVID-19. It uses POLYMOD age-mixing data [31] to quantify the age-mixing matrix and employs an equation [32] to quantify socially feasible types of mixing within and between deprivation deciles. We used analytical methods to analyse the equilibria and stability of the model. This study builds upon previous work by capturing how differences in both age and deprivation deciles, coupled with time-dependent testing capacity, influence disease transmission dynamics.

## 2. Mathematical Model Formulation

We consider an extended susceptible–exposed–infectious–recovered (SEIR) mathematical model of infectious diseases segregated into age groups *i* = 1, …, 21 and deprivation deciles *n* = 1, 2, …, 10. The model has eight epidemiological compartments: susceptible (S), exposed (E), asymptomatic (A), undetected symptomatic (J), detected symptomatic (I), hospitalised (H), recovered (R), and dead (D). Therefore, 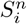 is the number of susceptible individuals in age group *i* and deprivation decile *n*; the corresponding notation is used for all other compartments. Essentially, the model segments individuals based on both age and socioeconomic factors (as indicated by a deprivation decile) to better understand and predict disease dynamics within these compartments. The total population by age and deprivation decile is defined as

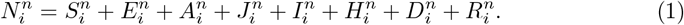

In this model, as shown in Figure 1, susceptible individuals are infected after coming into contact with an infectious individual at a rate 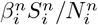, which is a function of social mixing, age mixing, differential susceptibility, infectivity by age, and relative transmission. The exposed individuals transition to other compartments after a latency period *σ* and a time-varying testing fraction *π*(*t*) divides them into either detected 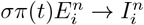 or undetected 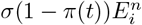. The undetected compartment was further subdivided into asymptomatic 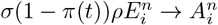 and undetected symptomatic 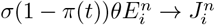 Asymptomatic individuals recover at a rate *γ*, and symptomatic individuals 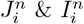 can recover or become hospitalised at rates *γ* and *δ*_*i*_, respectively. Hospitalised individuals can either recover or die from infection at rates *γ* and *d*_*i*_, respectively. We assumed that only hospitalised individuals died from the infection. We also assume that the test rate is a function of time because, at the start of an epidemic, the testing rate/capacity will be small compared to the later stages of the pandemic when testing is more readily available [36]. We defined the testing parameter *π*(*t*) using the generalised Richards model (GRM) (extended logistic function) [37] (*see supplementary material)*.

**Fig 1.**
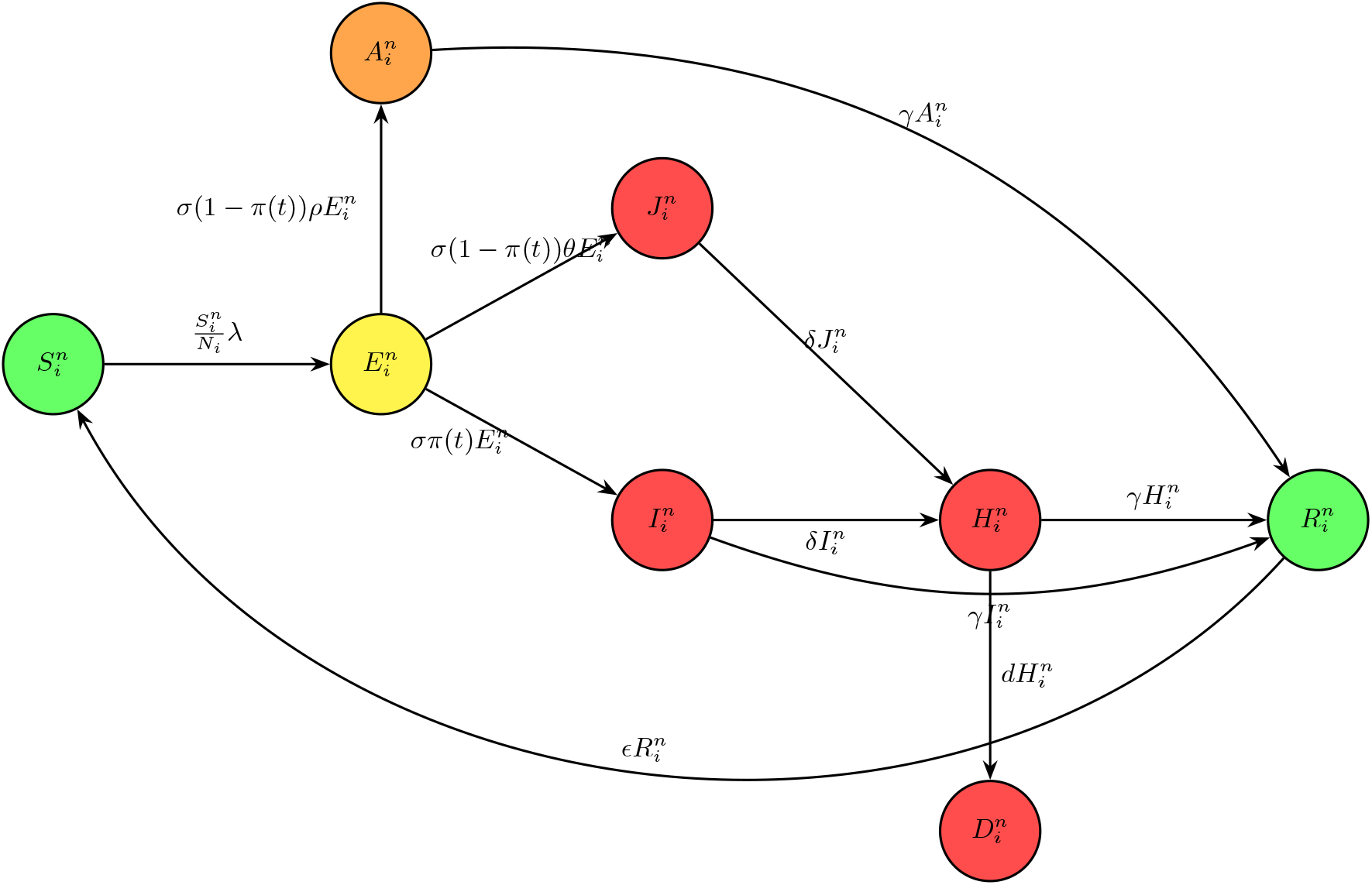
Schematic diagram of the epidemiological model of mixing within and between age groups and deprivation deciles. Susceptible individuals 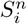 (green) acquire an infection at the rate 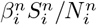 and enter the latent class 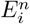 (yellow). At rate *σ*, the exposed hosts are detected as symptomatic with a time-varying testing rate *π*(*t*). Alternatively, at the rate (1 − *π*(*t*)), they can enter the asymptomatic class 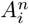 (orange) or the undetected symptomatic class 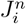 (red) in proportions *ρ* and *θ*. Asymptomatic, symptomatic, and hospitalised individuals recover to 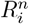 (green) at the rate *γ*. Detected and undetected symptomatic individuals may become hospitalised 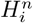 (red) at the rate *δi*.

### Key model assumptions

- The preferred mixing method (*ϵ* = 0.3 in equation 5) was adopted for interactions between deprivation deciles, and it was assumed that similar age mixing [31] occurs consistently across these deciles.
- The age–deprivation contact matrix is held constant over the simulated period. This omits temporal changes in behaviour (lockdowns, school schedules, holidays) or adaptive responses to rising cases.
- Births, natural deaths, ageing between groups, and migration are ignored over the simulated periods, so population sizes in each compartment change only via disease transitions.
- rate of recovery (*γ*), hospitalisation (*δ*_*i*_), and death (*d*_*i*_) are uniform across deciles. We assumed that deprivation affects only the contact rates.

### 2.1 Model Equations

The system of differential equations for the population in age group *i* and social group *n* is as follows:

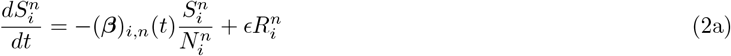

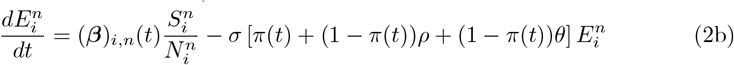

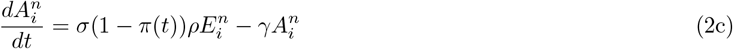

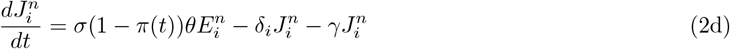

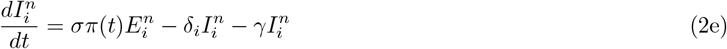

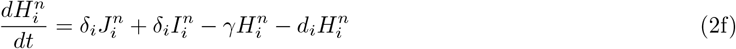

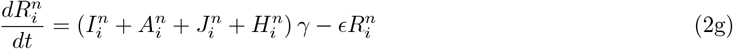

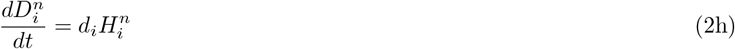

where *θ* = 1 − *ρ*, and *π*(*t*) + (1 − *π*(*t*))*ρ* + (1 − *π*(*t*))*θ* = 1. Here, (***β***)_*i,n*_(*t*) is defined as

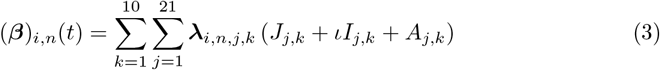

where ***λ***_*i,n,j,k*_ is the contact rate between individuals in age group *i* and social group *n* and those in age group *j* and social group *k* as shown in Figure 2. Additionally, ***λ***_*i,n,j,k*_ is the contact rate between individuals in age group *i* and social group *n* and those in age group *j* and social group *k*. Furthermore, ***λ***_*i,n,j,k*_ are elements of the block matrix ***λ***, which is a square matrix of size *n* × *n*, where *n* = 1, 2, …, 10 and each element is a matrix of size *i, j* = 1, 2, …, 21. In this model, we assume that only symptomatic and asymptomatic individuals **J**_*n,j*_, **I**_*n,j*_, and, **A**_*n,j*_ contribute directly to infection transmission, and that there is no transmission of infection in hospitals. In addition, we assume that asymptomatic individuals can transmit the disease without being aware of it, so we do not impose a modification factor for the transmission of asymptomatic individuals. In equation (3), we define

**Fig 2.**
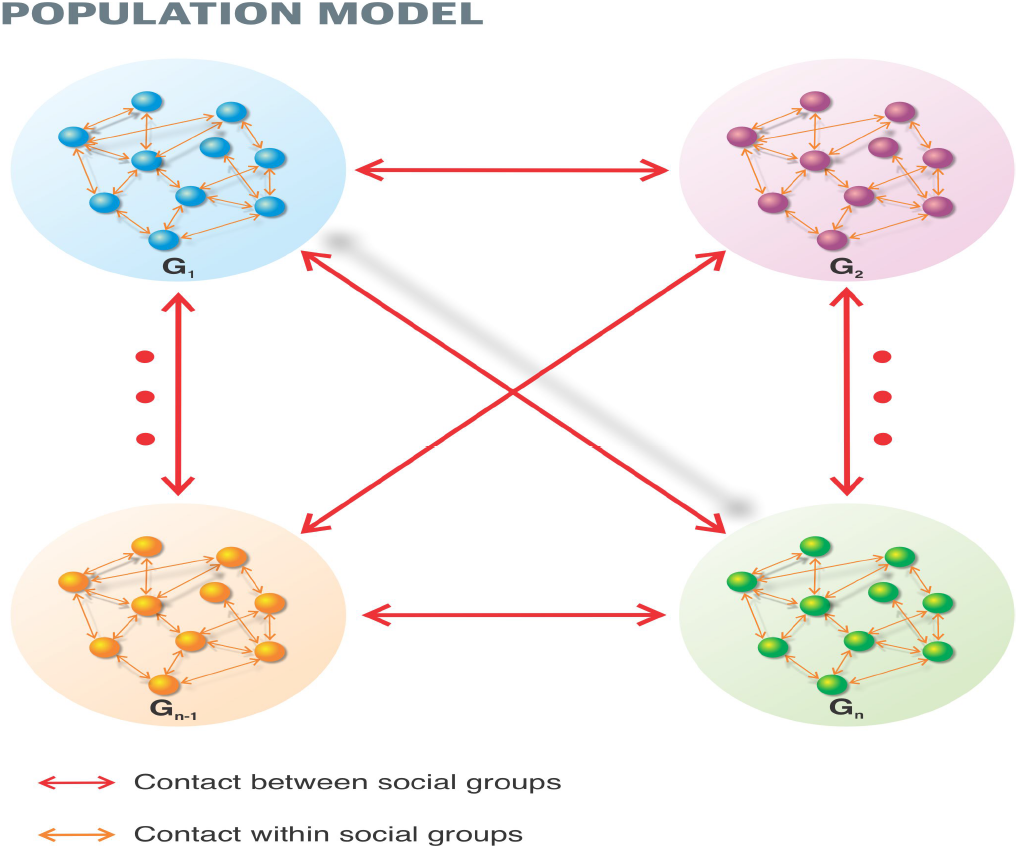
Population interactions by age and deprivation deciles. *G*_1_ … *G*_*n*_ are sociodemograhic groups which equate to deprivation deciles.

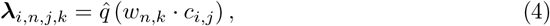

where **C** is the age contact mixing matrix from the study of Mossong *et al*. [31], 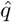 is the relative transmission rate, and the social interaction by deprivation decile is derived using the method of Hethcote [32], being defined as

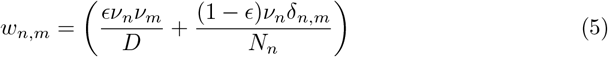

where (*δ*_*n,m*_ : *m* = *m is* 1, *& n* ≠ *m is* 0), *ϵ* ∈ [0, 1], and *ν*_*n*_ are the average number of contacts for each sociodemographic group. Here, *n* and *m* are the deprivation deciles, *N*_*n*_ is the total population of each decile in England, *D* = Σ_*n*=1_*ν*_*n*_*N*_*n*_ is the total number of contacts per unit time for everyone in the population.

### 2.2 Basic Qualitative Properties of Model 2

In this subsection, we prove that the solution to system (2) is mathematically and biologically relevant if and only if it is positive and bounded.

#### Theorem 1

(Positivity). *Let*

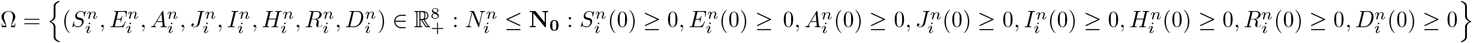*Then, the solution* 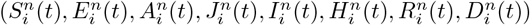 *of system* (*2) remains positive for all t >* 0.

*Proof*. The initial conditions 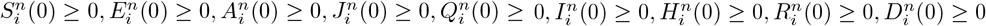 will be used to prove the positivity of the system (2). From equation (2a), we let

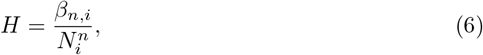

Equation (2a) can now be written as

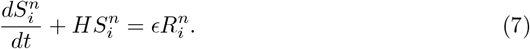

The integrating factor of equation (7) is given by

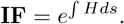

We multiply equation (7) by **IF**:

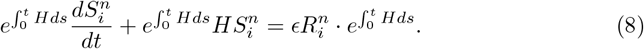

In equation (8), we observe that the LHS is the derivative of **IF** multiplied by 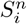 Hence, we can write equation (8) as:

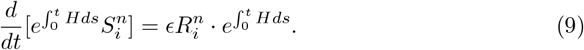

Taking the derivative of both sides, we obtain

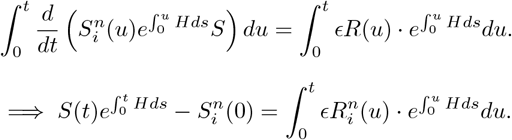

Further rearranging yields

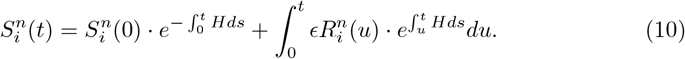

From the initial conditions, we know that 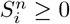 means that 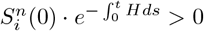 and all parameters are positive. That is, *ϵ >* 0, and hence,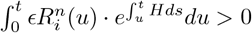. Therefore, we can conclude that 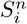 is positive for all *t >* 0. The same is true for 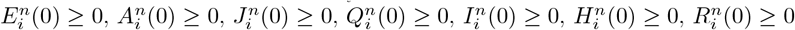 and 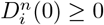.

#### Theorem 2

(boundedness). *All feasible solutions of the model system* (*2) are unifo mly bounded in a closed set* 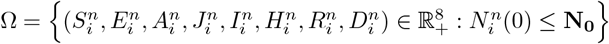.

*Proof*. At 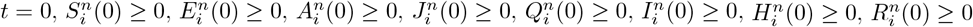, and 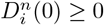. Considering the boundedness of the system (2) under the initial conditions such that

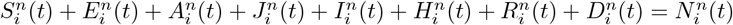, we have

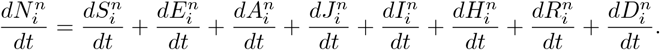

By taking the sum of the system of ordinary differential equations, we obtain

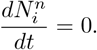

This implies that 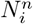 remains constant over time. By integrating both sides with respect to *t* and applying the initial condition 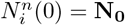, we obtain 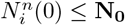 for all *t*. Hence, the feasible solution of the model system remains in the region 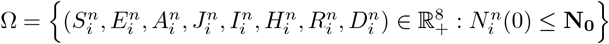.

## 3 Presentation of the analytical result

We analysed our model analytically to verify whether the model system (2) behaves consistently with epidemiological intuition.

### 3.1 Disease-Free Equilibrium and Basic Reproduction Number

The basic reproduction number is the average number of infected individuals in a completely susceptible population. Considering the system of equations (2a–2h) and the total population in equation (1), the infected compartments are 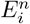 (exposed), 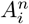 (asymptomatic), 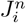 (undetected symptomatic), and 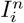 (detected symptomatic). The compartments 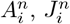 and 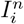 contribute directly to the force of infection, whereas 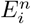 is the point at which a susceptible individual comes into contact with an infected individual. In the disease-free equilibrium (DFE), 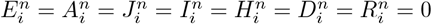 and 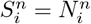.

We solve for *R*_0_ using the next-generation matrix approach and then determine the dominant eigenvalue of (𝒯 𝒱 ^−1^). We divide the system into disease transmission (𝒯) and disease transition (𝒱 ^−1^), as follows:

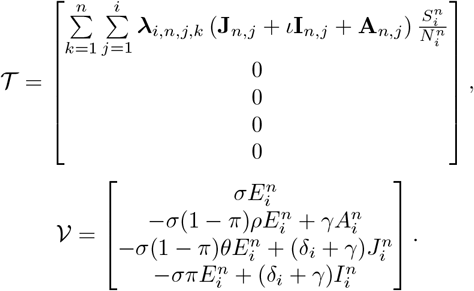

The Jacobian of the two systems at the DFE and for *θ* = 1 − *ρ* is calculated as

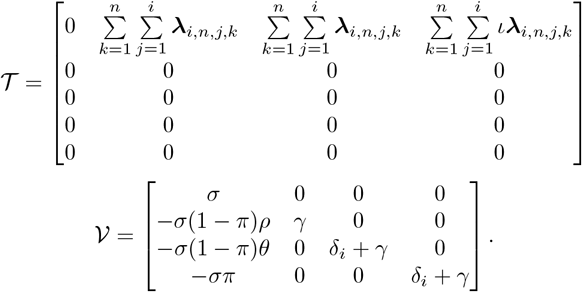

The inverse of 𝒱 is given by

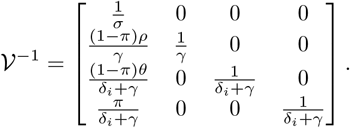

Hence, we can compute 𝒯 𝒱 ^−1^ for one group as

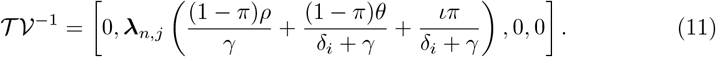

For multiple groups, we construct a full matrix

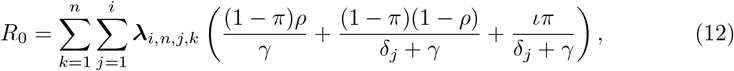

From equation (12), *R*_0_ is largely dependent on the mixing matrix (which is the combination of age and deprivation mixing).

### 3.2 Stability of the Disease-Free Equilibrium

#### Theorem 3

(local stability). *The DFE of system* (*2) is locally asymptotically stable* (*LAS) if R*_0_ *<* 1 *and unstable if R*_0_ *>* 1.

*Proof*. To demonstrate the local stability of our model, we linearise the system by finding the Jacobian at the DFE. The Jacobian matrix at the DFE, where 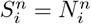 and all other compartments are zero, is an 8 × 8 matrix

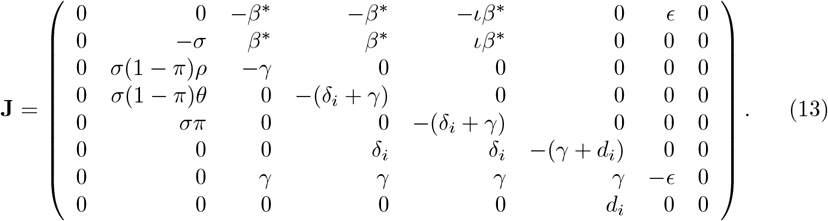

For simplicity, let 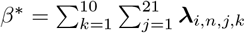. In this Jacobian matrix, the only nonzero entries in the first and last rows are *ϵ* and *d*_*i*_, representing the rates at which individuals lose their immunity to become susceptible and die from a severe infection, respectively.

These rates are negligible at DFE, as there are no infections, recoveries, or disease-related deaths. This implies that the impact of susceptibility loss due to infection exposure and infection-related death on the dynamics of other compartments is minimal. Hence, we focus on disease dynamics by removing the 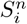 and 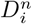 compartments and reducing the system to six compartments: 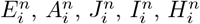, and 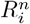. The differential equations for these compartments at DFE are

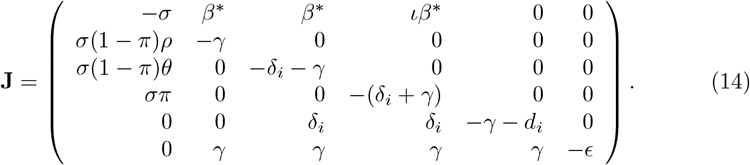

We obtain the characteristic equation by setting |*J* − *λI*| to zero:

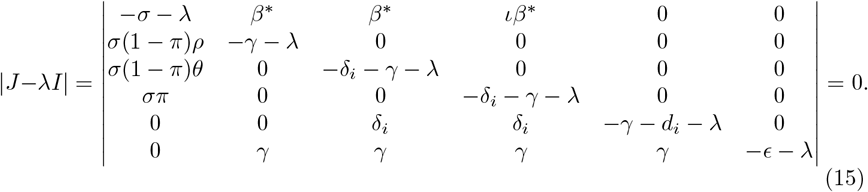

Hence, the coefficient of the characteristic polynomial is given by

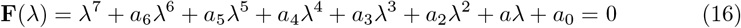

where

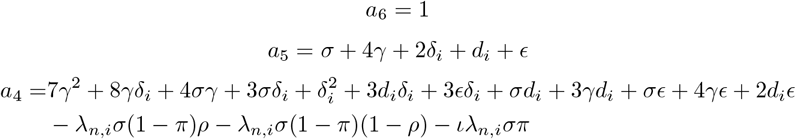

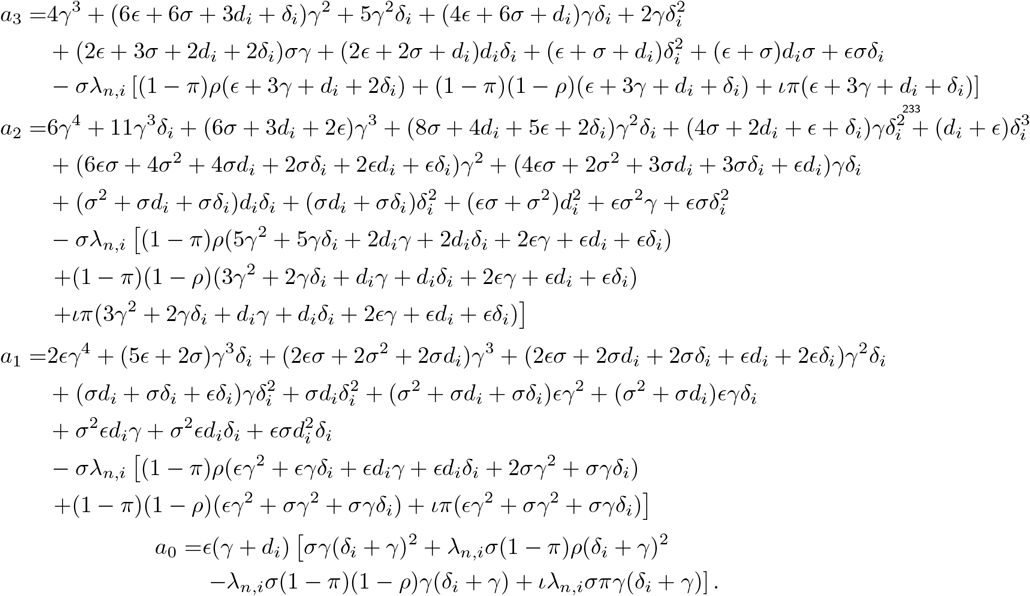

According to the Routh–Hurwitz condition, for *R*_0_ *<* 1, the DFE is locally asymptotically stable if *a*_6_ *>* 0, *a*_5_ *>* 0, *a*_4_ *>* 0, *a*_3_ *>* 0, *a*_2_ *>* 0, *a*_1_ *>* 0, and *a*_0_ *>* 0. That is,

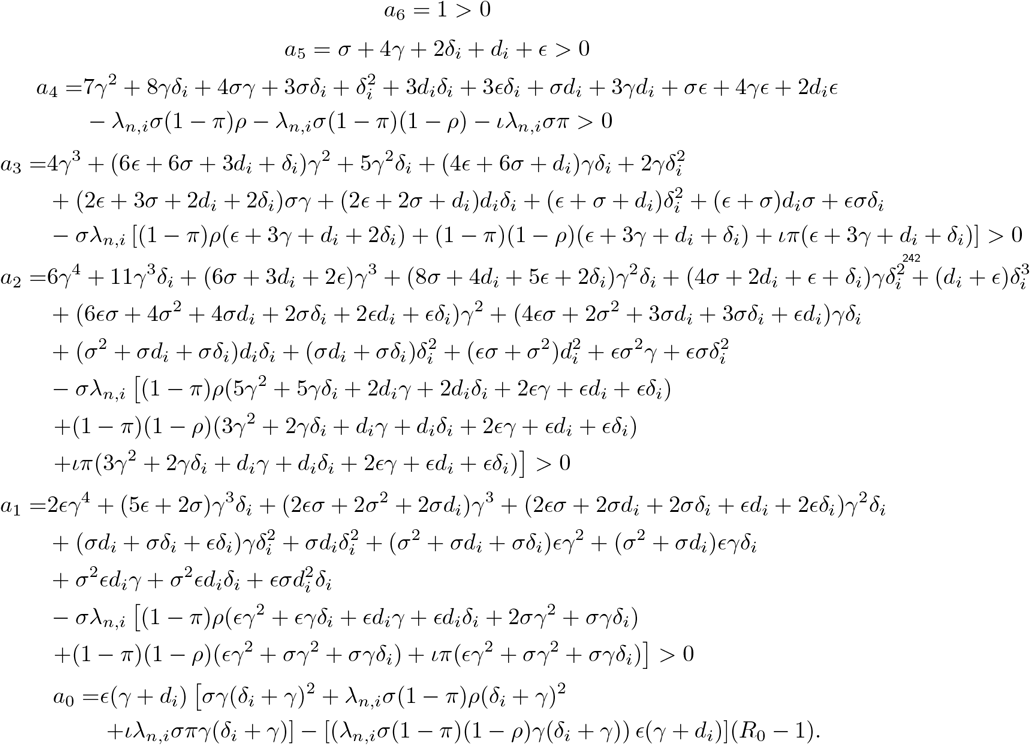

Thus, *a*_0_ *>* 0 if *R*_0_ *<* 1. Since all the coefficients are positive, we conclude that for *R*_0_ *<* 1, the DFE point of our model is locally stable.

#### Theorem 4

(global stability). *The system* (*2) is globally asymptotically stable at the DFE if R*_0_ *<* 1 *and unstable if R*_0_ *>* 1.

*Proof*. Let us consider the Lyapunov function *V* of the form

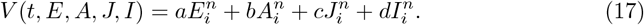

Its derivative is

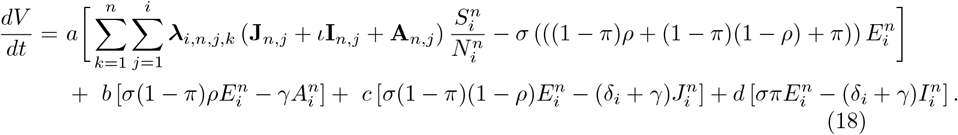

Rearranging such that like terms are collected together, and noting that at DFE, 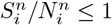, we have

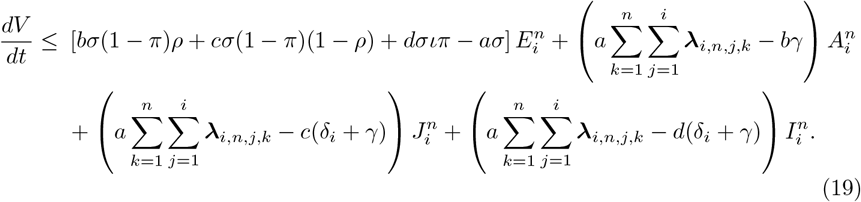

Let 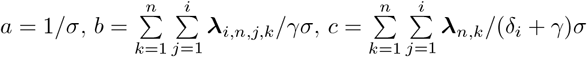 and

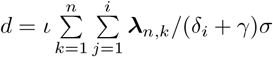. Substituting these into equation (19) leads to

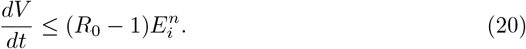

Hence,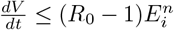; it is important to note that 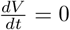 only at DFE. From LaSalle’s invariance principle, it follows that all solutions of equation (2) converge to the DFE as *t* → ∞ whenever *R*_0_ *<* 1. The DFE is globally asymptotically stable when *R*_0_ *<* 1.

### 3.3 Existence of an Endemic Equilibrium Point

We calculate the endemic equilibrium by equating the system (2) to zero; then, we obtain 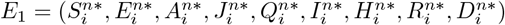, such that

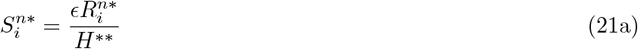

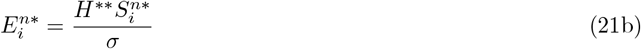

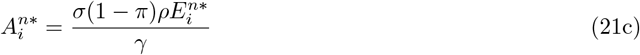

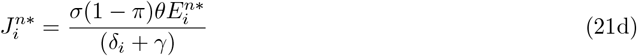

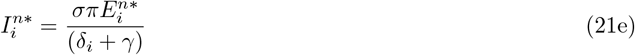

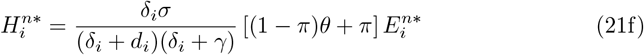

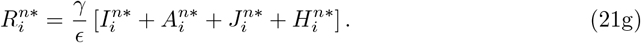

From equation (6), we define

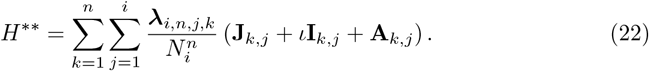

At endemic equilibrium, we know that 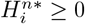 and that all our parameters are positive, so *d*_*i*_*≠* 0. We can conclude that 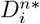 depends on 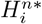.

#### 3.3.1 Uniqueness of the Endemic Equilibrium Point

##### Theorem 5.

*An endemic equilibrium is unique iff R*_0_ *>* 1.

*Proof*. If we rewrite equations (21c–21g) in terms of equation (21b), we have

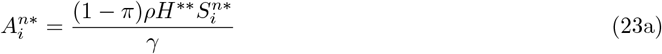

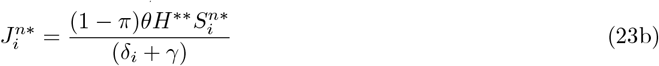

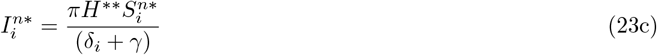

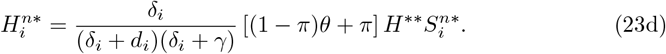

Substituting 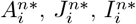, and 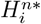 from equations (23a–23d) into (21g), we obtain

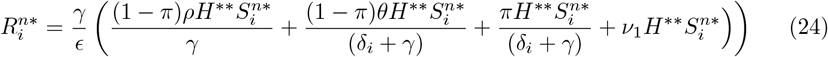

where

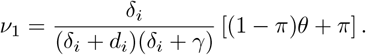

Equation (22) implies that

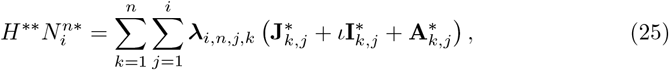

which implies that

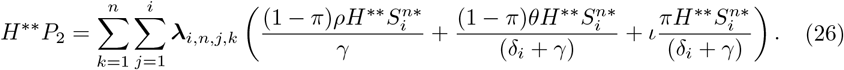

*P*_2_ is derived from 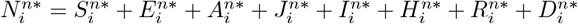. Dividing both sides of equation (26) by 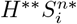, we have

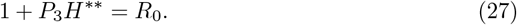

Solving for *H*^∗∗^ in equation (27), we have

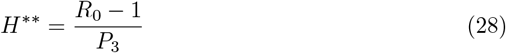

and *P*_3_ *>* 0. Here, *P*_3_ is defined as

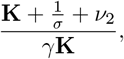

Where 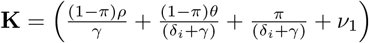, and

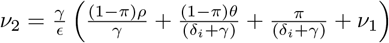. All parameters are positive; hence, *γ >* 0 and **K** *>* 0, so *γ***K** *>* 0. Therefore, *H*^∗∗^ *>* 0 iff *R*_0_ *>* 1. Therefore, a unique endemic equilibrium exists.

### 3.4 Global Stability of the Endemic Equilibrium

#### Theorem 6

(global stability of endemic equilibrium). *The endemic equilibrium E*_1_ *of the model system* (*2) is globally asymptotically stable if R*_0_ *>* 1.

*Proof*. To prove global stability, we used the Lyapunov function method. Consider the Lyapunov function of the form

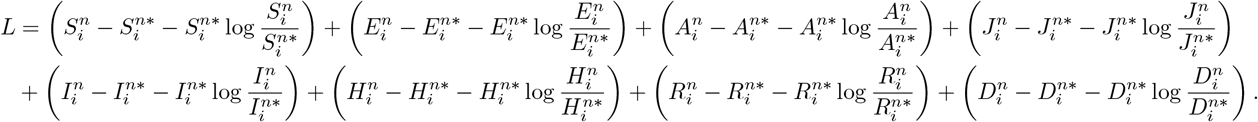

Taking the derivative of *L*, we obtain

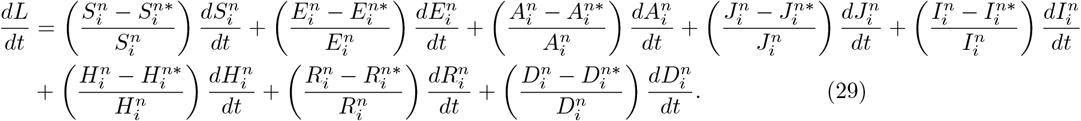

Substituting the value of the derivatives into (29) yields

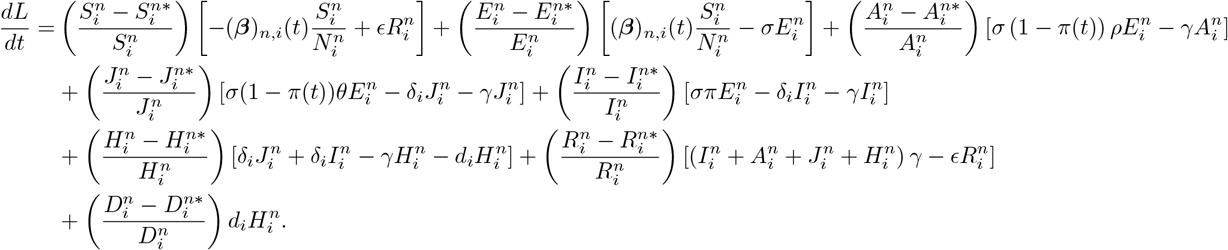

By substituting (*β*)_*n,i*_, we obtain

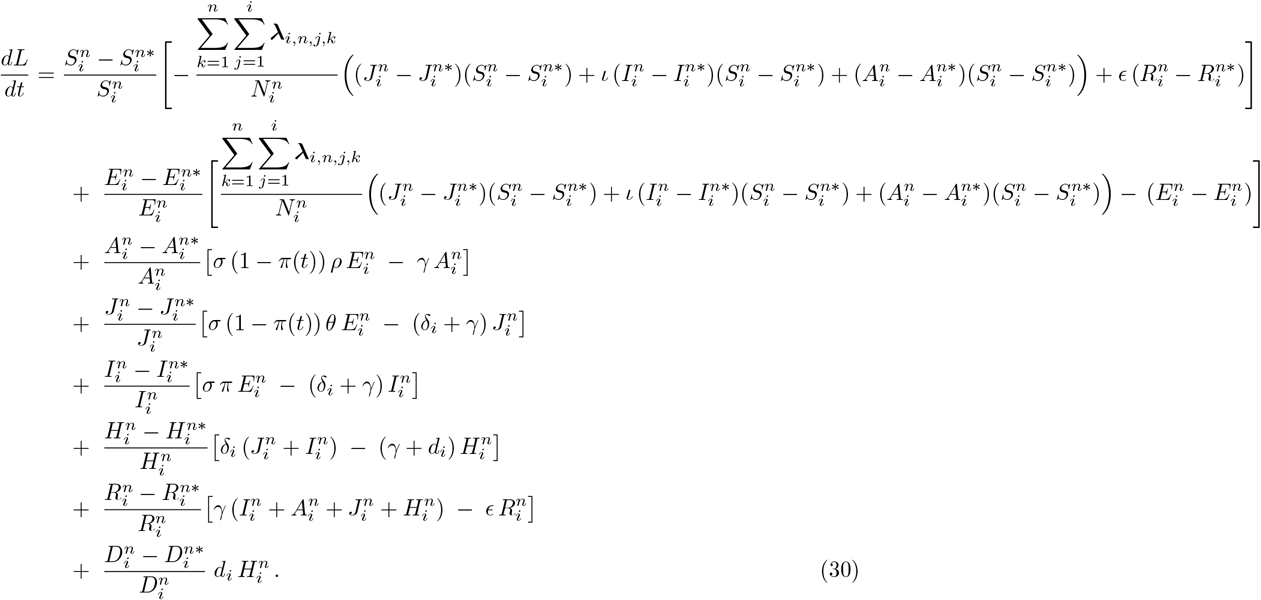

After some algebra, it is possible to split equation (30) into positive and negative parts:

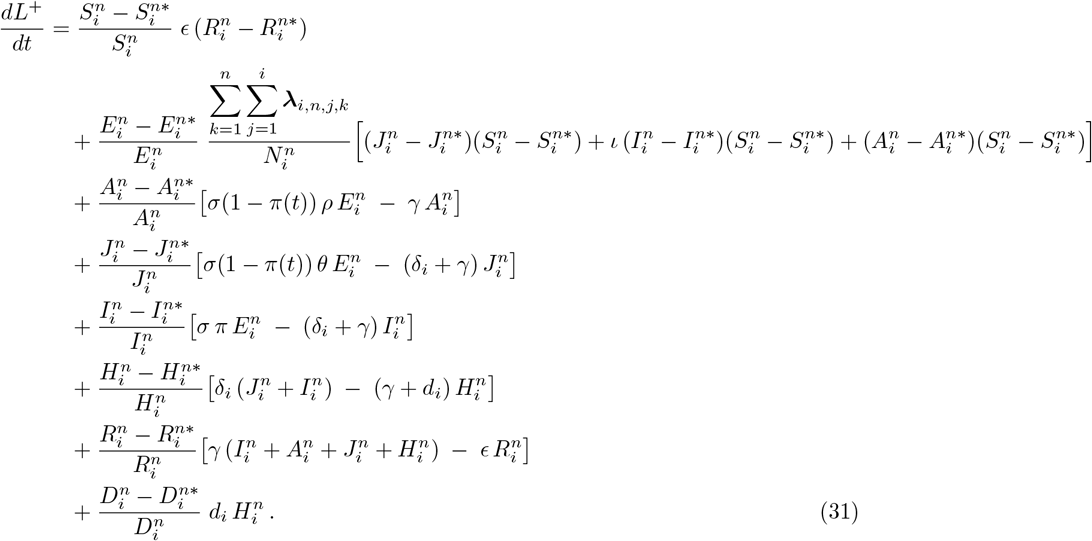

Taking the positive terms, we have

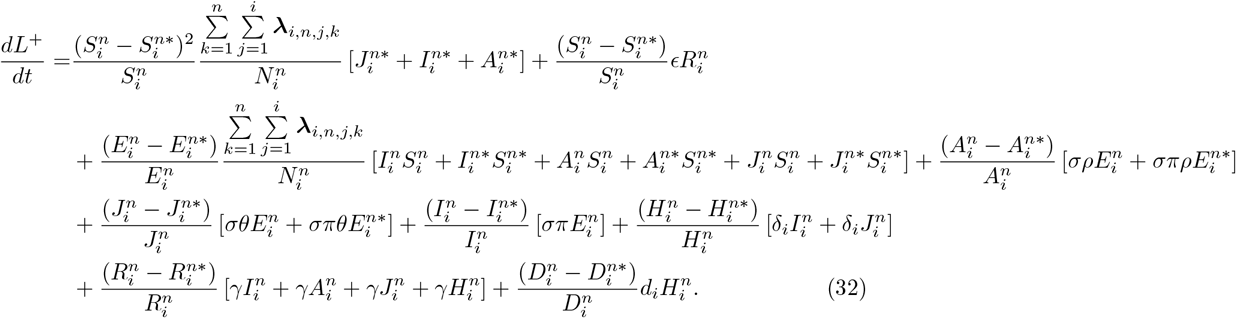

Taking the negative terms, we have

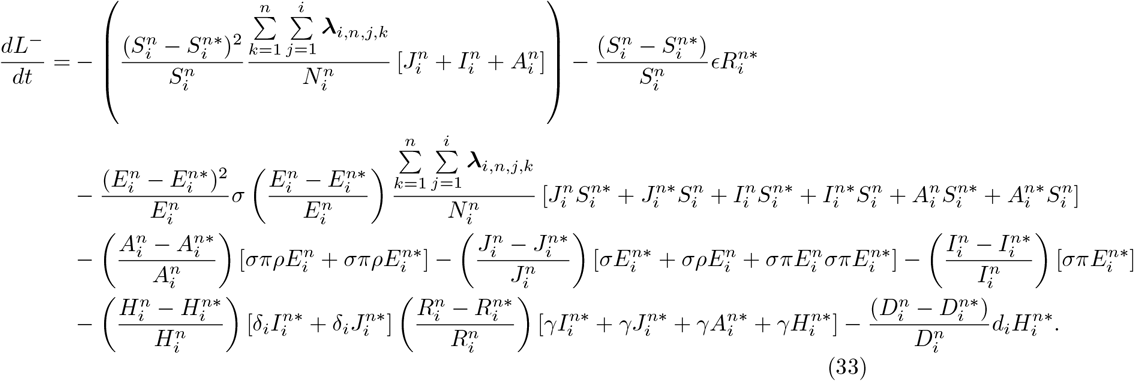

We can write 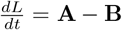, where **A** represents the positive terms (32) and **B** the negative terms (33). Thus, if **A** *<* **B**, then we obtain that 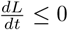 iff 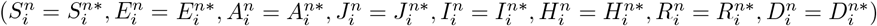. Therefore, the largest compact invariant set in 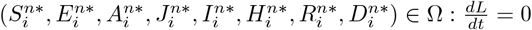 is the singleton set, *E*_*eff*_, where *E*_*eff*_ is the endemic equilibrium of the system.

According to LaSalle’s invariance principle, this implies that *E*_1_ is globally asymptotically stable in Ω if **A** *<* **B**.

### 3.5 Sensitivity Analysis

We performed a sensitivity analysis to study how the uncertainty in the model output could be predicted by the uncertainty in the model input. This analysis helps the mathematical modeller validate their assumptions and improve trust in the results of the model. The normalised sensitivity index for each parameter *X* within *R*_0_ is defined as

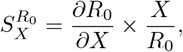

which was used in studies by Wodajo *et al*. [38] and Al-arydah [39]. Parameters with a high sensitivity index have a greater influence on the *R*_0_ value. The sign of the index indicates whether the parameter influences *R*_0_ positively or negatively. From Figure 3, it can be seen that the contact rate, proportion of asymptomatic individuals in the population, testing rate, recovery rate, and rate of hospitalisation are the key parameters that influence the reproduction number, whereas the sensitivity of the test and average number of days for the test to be reported have a small influence.

**Fig 3.**
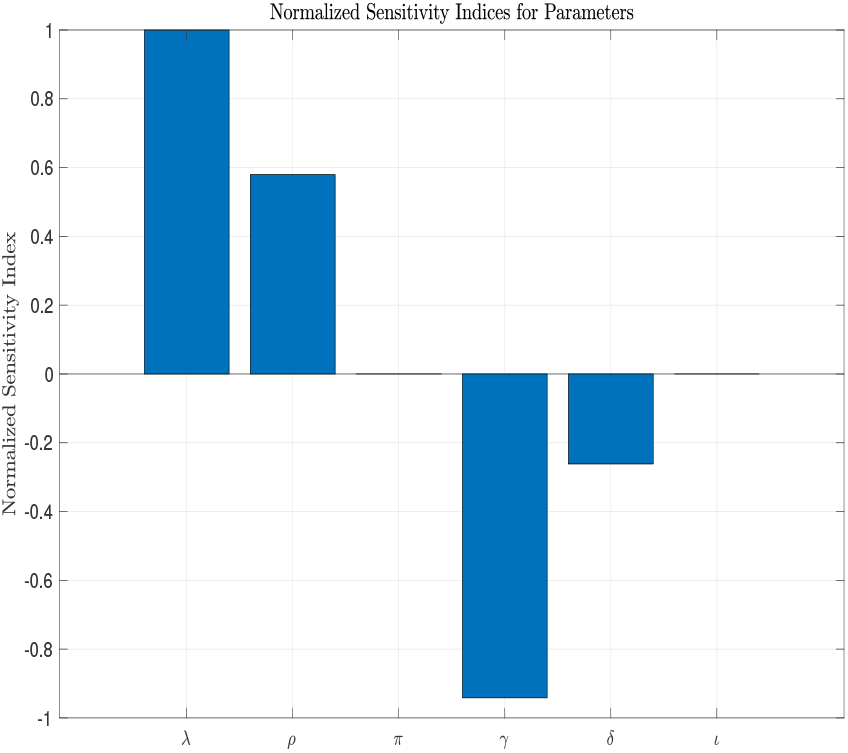
Bar chart of normalised sensitivity indices for key model parameters affecting the basic reproduction number *R*_0_. Each bar represents the relative impact of changes in the model parameters, with positive values indicating parameters that increase *R*_0_ and negative values indicating those that decrease it. High sensitivity values imply that small changes in these parameters can significantly affect the disease transmission. Key parameters include the contact rate *λ*, recovery rate *γ*, and hospitalisation rate *δ*.

**Fig 4.**
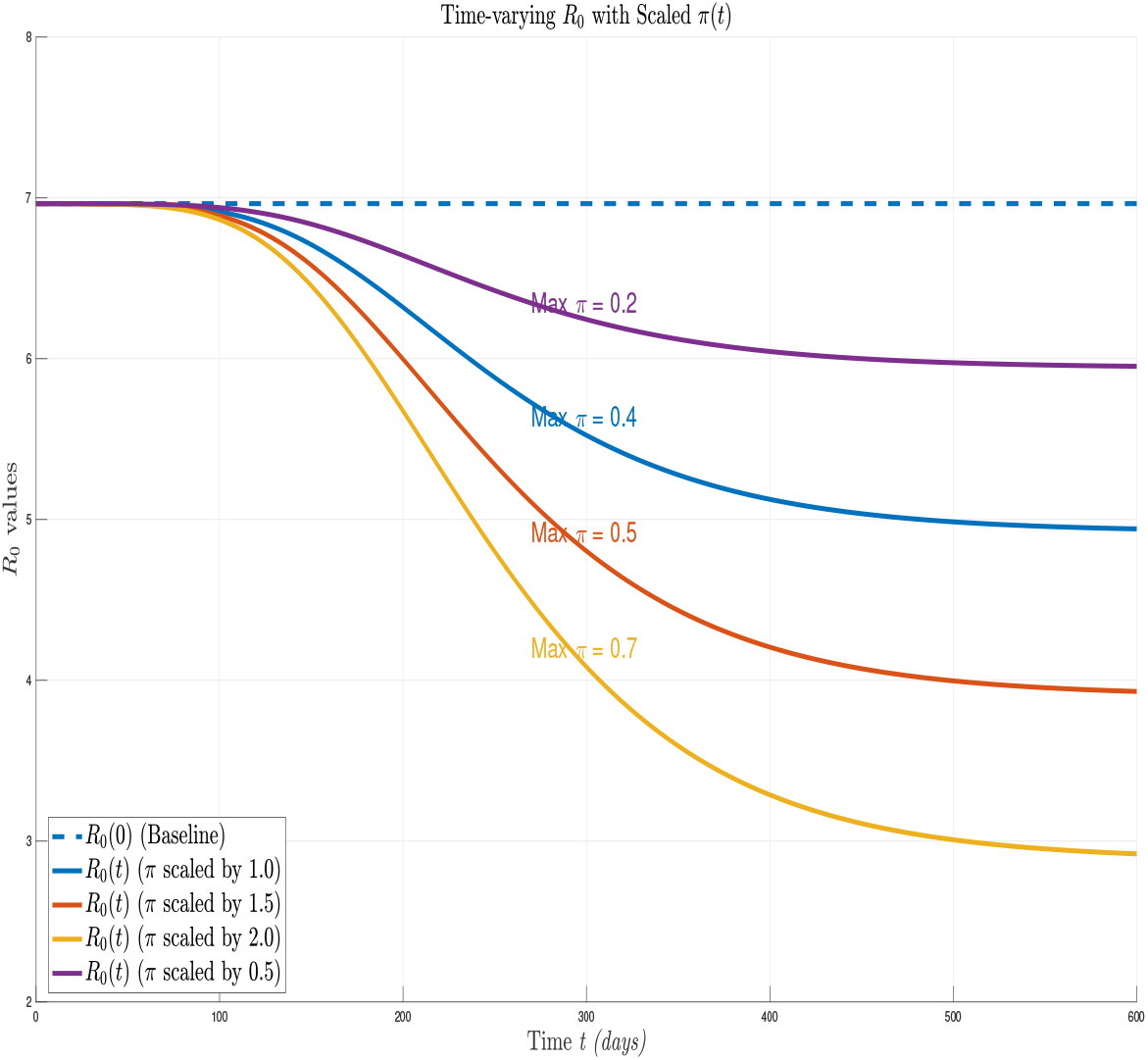
Time-varying basic reproduction number *R*_0_(*t*) under different testing rate trajectories *π*(*t*). The dashed horizontal line marks thse initial reproduction number *R*_0_(0) *≈* 7.0. Solid curves show how *R*_0_(*t*) declines as the cumulative testing capacity increases according to a logistic “generalized Richard” curve scaled by factors 0.5, 1.0, 1.5, and 2.0 (yielding maximum proportions of the testpopulation of 0.2, 0.4, 0.5, and 0.7, respectively). Increased testing reduces the proportion of undetected (and thus fully transmissible) infections, lowering *R*_0_, but—even at the highest feasible testing scale—cannot on its own drive *R*_0_ below 1.

The sensitivity indices were expressed as follows:

1. The sensitivity index for *λ*_*i,n,j,k*_ is

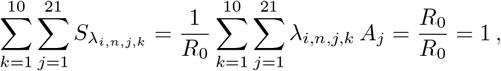

where 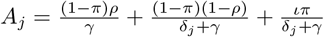. This indicates that the normalised sensitivity index of *λ*_*i,n,j,k*_ is a matrix of ones.
2. The sensitivity index for *γ* is

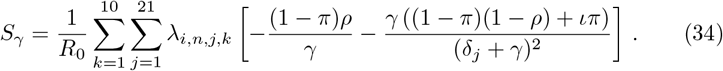
3. The sensitivity index for *ρ* is

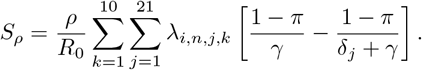
4. The sensitivity index for *δ*_*j*_ is

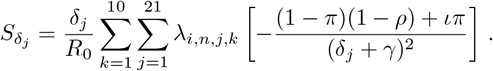

However, we can compute the sensitivity index specific to each *j* by summing only over the index *k*. The normalised sensitivity index for *δ*_*j*_ is given by

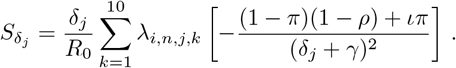
5. The sensitivity index for *π* is

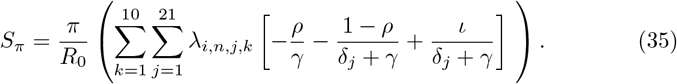
6. The sensitivity index for *ι* is

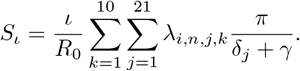

## 4 Numerical Analysis

We numerically analysed the sensitivity indices associated with each parameter. Numerical analysis was performed in MATLAB using the built-in ode45 solver with the parameter values listed in Table 1. The initial conditions for each age group and deprivation decile were set such that the entire population was susceptible, except for a small seed of latent infections 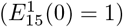 to initiate spread. The parameter values were obtained from existing literature, except where otherwise stated. A sensitivity analysis was performed using the sensitivity index method and numerically by perturbing some of the parameters to observe their impact on the system dynamics, as presented in Figure 5. Under the baseline mixing patterns, relative transmission rate 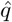, and parameter values, the next-generation matrix approach yields a basic reproduction number of *R*_0_ = 3.34. However, when we reduce the relative transmission rate 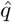 by 80%, the *R*_0_ values decrease below 1, indicating that a reduction in the transmission rate, which directly affects the mixing and contact patterns, is sufficient to take *R*_0_ below the threshold for sustained spread. Finally, Figures 6 and 7 illustrate the time courses of all compartments (susceptible, exposed, asymptomatic, infectious, hospitalised, recovered) aggregated over age for each of the ten deprivation deciles. It can be seen that the more deprived groups peaked earlier and attained higher burdens of infection and hospitalisation.

**Table 1.** Parameter descriptions and values.

| Parameter | Description | Value and Source |
| --- | --- | --- |
| $c_{i,j}$ | Contact matrix by age | [31] |
| $\iota$ | Modification factor by which confirmed and reported individuals can transmit an infection. | 0.6 (assumed) |
| $\rho$ | Proportion of population who are asymptomatic | 0.6 |
| $\pi(t)$ | Time-dependent fraction of exposures reported | fitted |
| $\hat{q}$ | Relative transmission rate | 2.5 (assumed) |
| $\gamma$ | Recovery rate | 1/14 days<sup>-1</sup> [34] |
| $\delta_i$ | Rate at which symptomatic or positive quarantine individuals become sick and require hospital care, which varies by age | [33] |
| $d_i$ | Rate at which hospitalised individuals become fatally sick, which varies by age | [33] |
| $\epsilon$ | Probability of waning immunity | 2/100 days<sup>-1</sup> [35] |
| $a_i$ | Infectivity by age | [33] |
| $h_i$ | Susceptibility by age | [33] |

**Fig 5.**
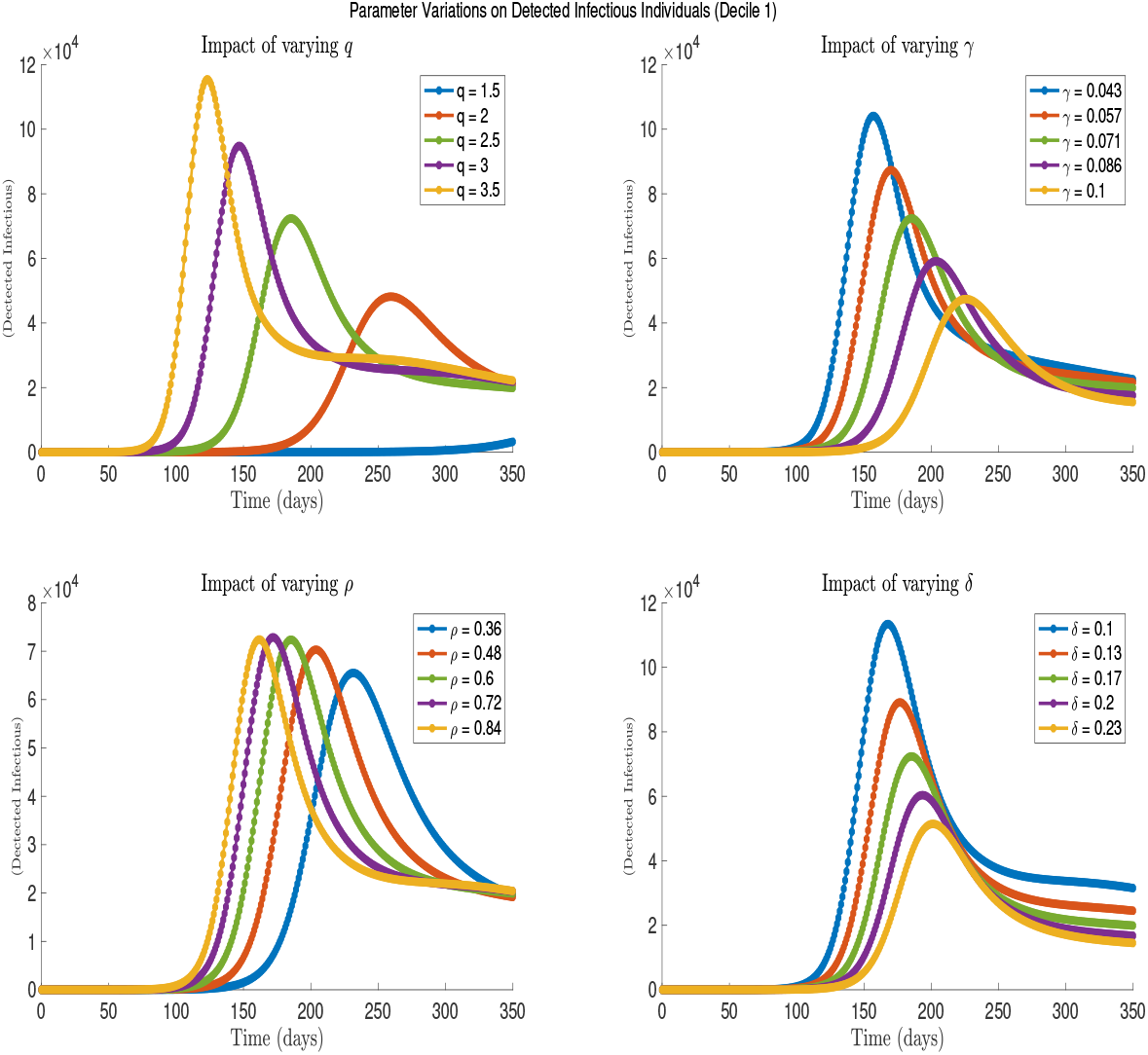
Sensitivity of infection dynamics in the most-deprived decile (Decile 1) when *R*_0_ *>* 1. Each panel shows the time series of detected infectious individuals under *±* 20% variation from baseline values in some key parameters: relative transmission rate 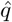, proportion of asymptomatic *ρ*, recovery rate *γ*, and the rate of hospitalisation *δ*. Changes in these parameters can have a significant impact on the peak of infection.

**Fig 6.**
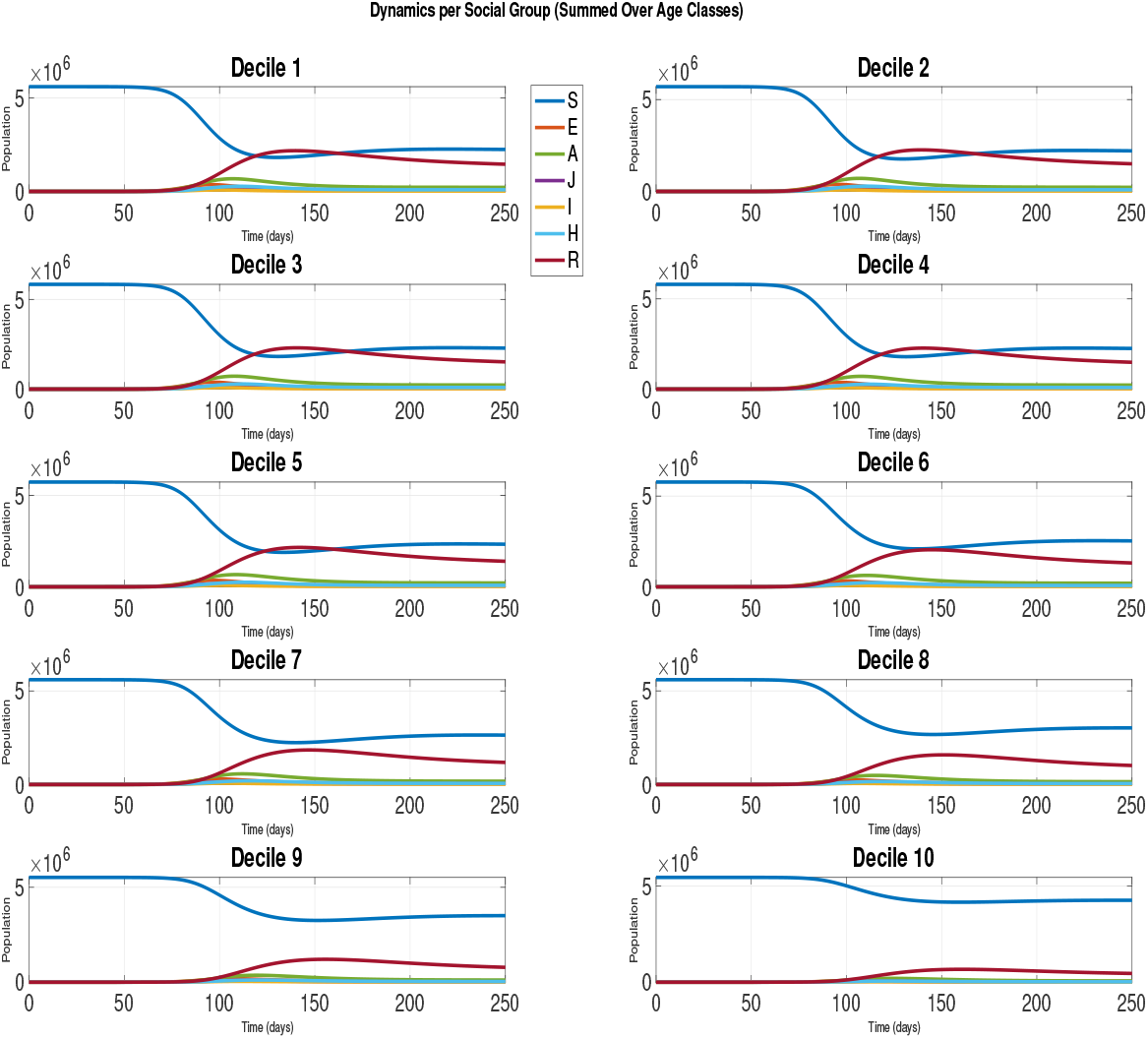
Time series of all epidemiological compartments aggregated over age for each deprivation decile. Panels1–10 correspond to deciles1 (most deprived) to10 (least deprived). For each, the curves show the susceptible, exposed, asymptomatic, symptomatic (detected and undetected), hospitalised, and recovered populations over 150 days. More deprived deciles peak earlier and at higher magnitudes in infectious and hospitalised cases, reflecting denser contact patterns.

**Fig 7.**
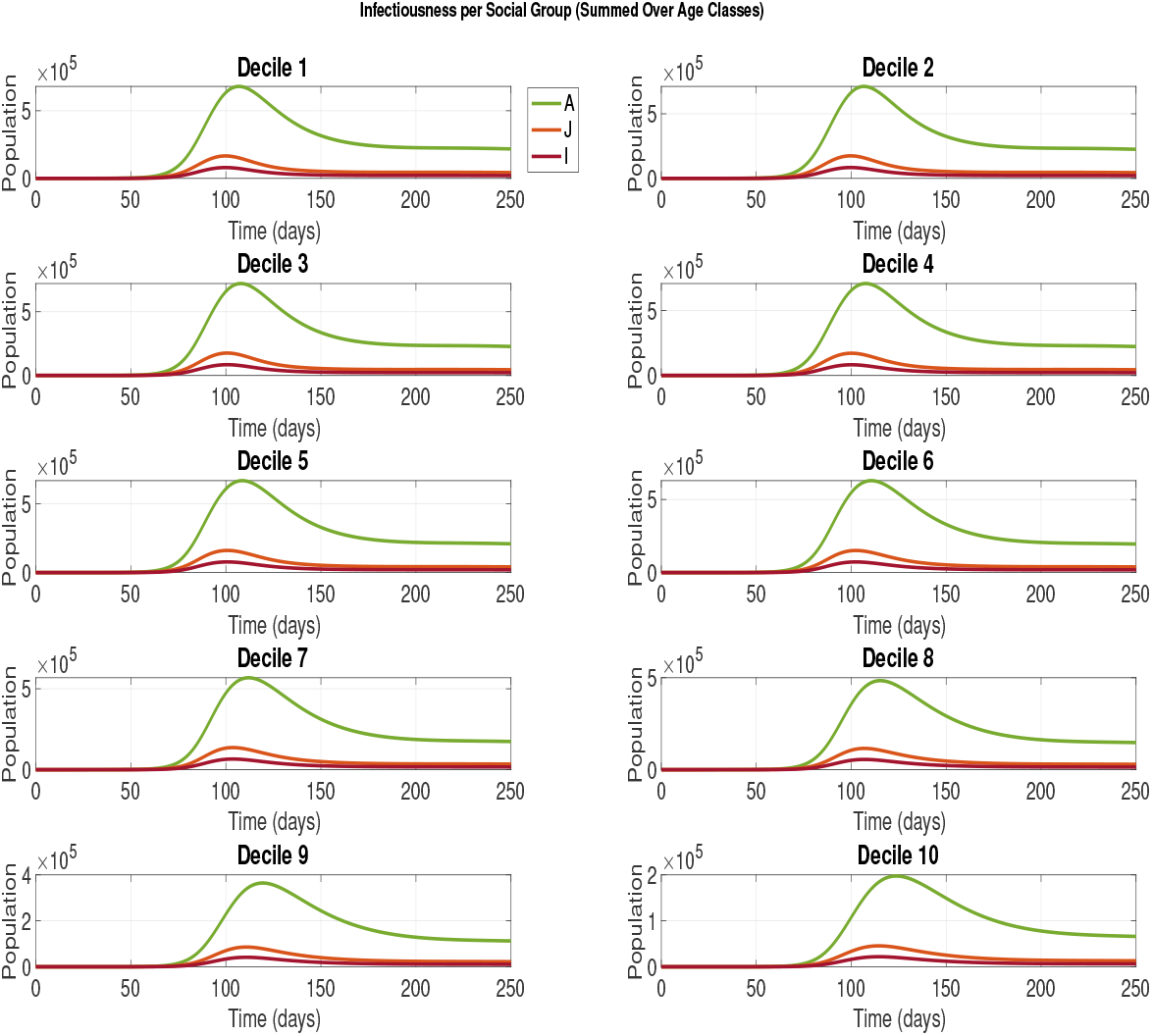
Infection trajectories by deprivation decile, aggregated over age groups. For each decile (1=most deprived… 10=least deprived), the curves show asymptomatic (green), detected symptomatic (red), and undetected symptomatic (orange), populations over 250 days. More-deprived deciles peak earlier and reach higher burdens in the infectious compartments.

### 4.1 Simulation of model (2) at *R*_0_ *>* 1

In Figure 6, we plotted the full time series of the model’s eight compartments (susceptible, exposed, asymptomatic, undetected, and detected symptomatic, hospitalised, recovered, and deceased) aggregated over age for each deprivation decile using the baseline parameter set (Table 1). Deciles1–10 are shown in separate panels, highlighting that the most-deprived groups experience earlier and larger infection peaks. In Figure 7, we focus on the breakdown of the infectious compartments, again summed over age within each decile, and using the baseline parameter set (Table 1).

### 4.2 Pairwise variation of key epidemiological parameters

In Figure 8, we hold the age–deprivation contact matrix fixed and compute *R*_0_ across a *±*20% grid of the relative transmission rate 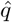, proportion of asymptomatic individuals *ρ*, recovery rate *γ*, and hospitalisation rate *δ*. Darker shading indicates a higher *R*_0_. We see that *R*_0_ is most sensitive to 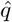 and *γ*; reducing transmission or shortening the infectious duration yields the greatest decline in *R*_0_. Figure 9 repeats these comparisons but now varies the overall contact-mixing scale *λ* vs. each of *ρ, γ*, and *δ* in the same six-panel layout. We deduce that interventions aimed at minimising contact, in conjunction with key measures such as improving recovery through treatment, are crucial to effectively manage the spread of the disease.

**Fig 8.**
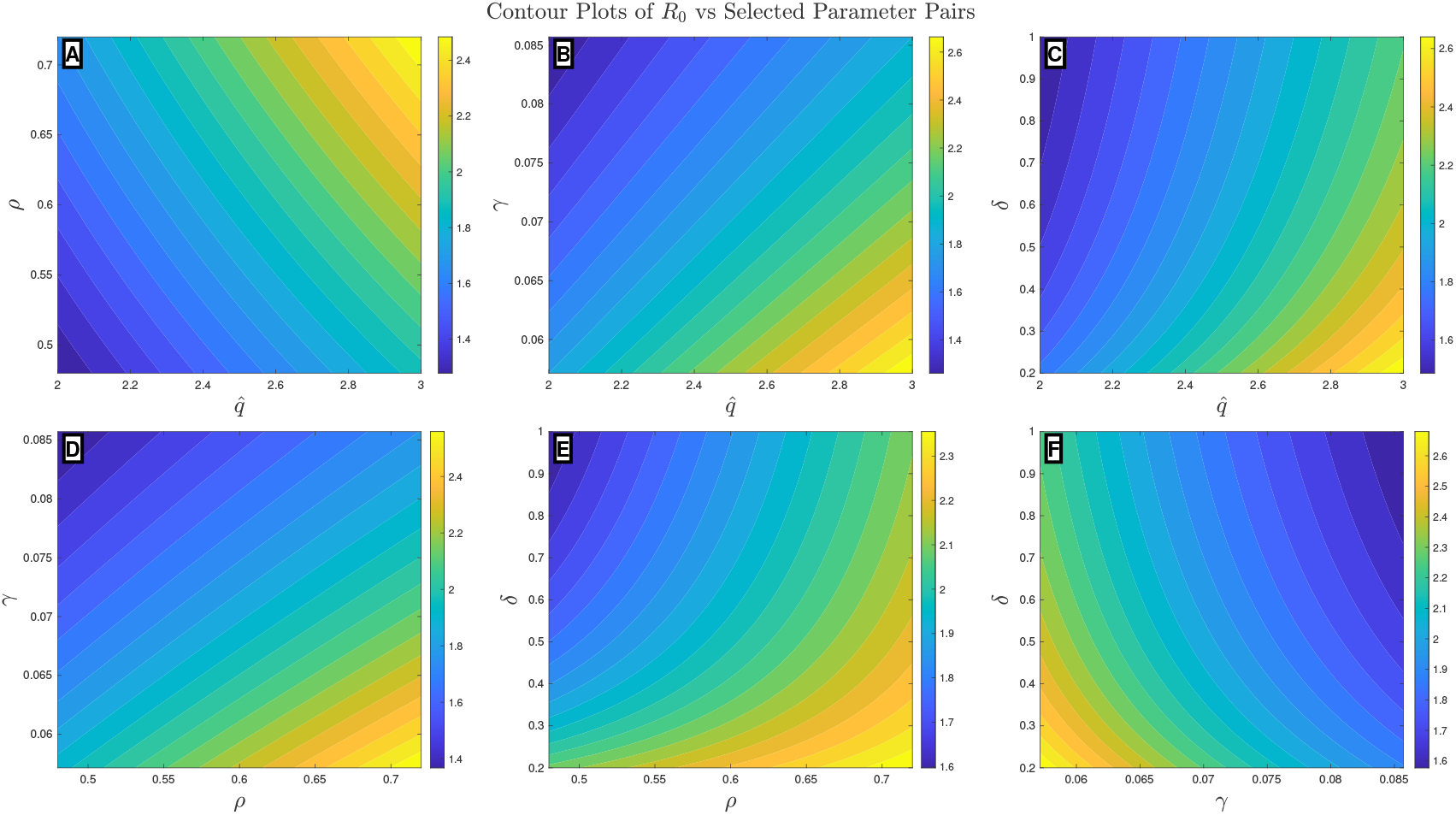
Contour plots illustrating the sensitivity of the basic reproduction number *R*_0_ to pairwise variations in the key epidemiological parameters. Each panel shows *R*_0_ computed over a *±*20 % grid around the baseline values of the two parameters, with all other parameters fixed. *δ*_*i*_ the rate of hospitalisation by age group is scaled to *δ* by the value [0.2, 1]. The pairs of parameters are (A) relative transmission rate 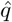 vs asymptomatic fraction *ρ*, (B) 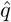 vs recovery rate *γ*, (C) 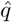 vs hospitalisation rate *δ*, (D) *ρ* vs *γ*, (E) *ρ* vs *δ*, and (F) *γ* vs. *δ*. Shading denotes the magnitude of *R*_0_ according to the adjacent colour bar.

**Fig 9.**
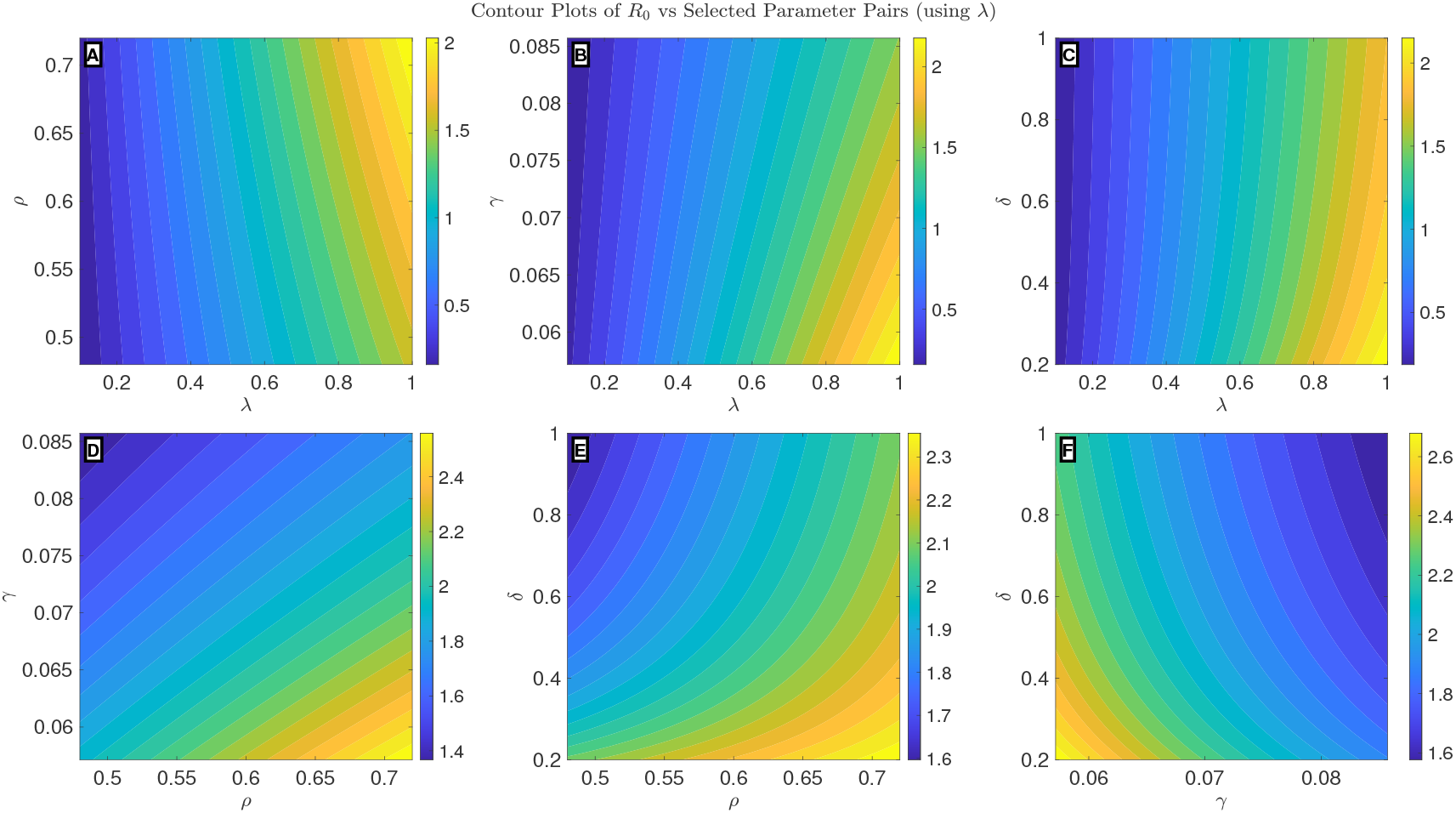
Contour plots illustrating the sensitivity of the basic reproduction number *R*_0_ to pairwise variations in key epidemiological parameters. Each panel shows *R*_0_ computed over a *±*20 % grid around the baseline values of the two parameters, with all other parameters fixed. The contact mixing matrix is scaled between [0.1, 1] and *δ*_*i*_ the rate of hospitalisation by age group is scaled to *δ* by the value [0.2, 1]. The pairs of parameters are (A) scaled contact matrix *λ* vs asymptomatic fraction *ρ*, (B) *λ* vs recovery rate *γ*, (C) *λ* vs hospitalisation rate *δ*, (D) *ρ* vs *γ*, (E) *ρ* vs *δ*, and (F) *γ* vs *δ*. The shading denotes the magnitude of *R*_0_ according to the adjacent colour bar.

## 5 Discussion and Conclusion

This study presented a compartmental model that stratifies the population by 21 age bands and 10 deprivation deciles, extending the classic SEIR framework to eight compartments: susceptible, exposed, asymptomatic, undetected symptomatic, detected symptomatic, hospitalised, recovered, and dead. This layered structure allowed us to capture two critical factors: age-dependent contact patterns (using the POLYMOD matrix) and socioeconomic mixing (using a deprivation-decile interaction matrix). By combining the two factors into a single “who-acquires-infection-from-whom” block matrix ***λ***_***i***,***n***,***j***,***k***_, we derived an expression for the basic reproduction number, as presented in equation (12), whose dominant eigenvalue quantifies the average number of secondary cases in a fully susceptible, stratified population. By examining the maximum eigenvalues of each block, we further decomposed the transmission contributions within and between deprivation deciles.

Analytically, we demonstrated the positivity and boundedness of all compartments, ensuring the absence of negative populations or unbounded growth. Using the next-generation matrix method, we showed that when *R*_0_ *<* 1, the DFE is asymptotically stable both locally and globally. This guarantees eventual epidemic extinction, regardless of the initial conditions. Additionally, we show that a unique endemic equilibrium exists and is globally asymptotically stable when *R*_0_ *>* 1. This confirms sustained transmission in the absence of interventions.

Sensitivity analysis, both analytical and numerical, revealed the parameter that most strongly drives the value of *R*_0_. The contact rate ***λ***_***i***,***n***,***j***,***k***_ has a normalised sensitivity index of 1, as shown in Figure 3. This implies that a 1% increase in the overall mixing directly translates to a 1% increase in the value of *R*_0_. The fraction of asymptomatic individuals *ρ*, recovery rate *γ*, and hospitalisation rate *δ* (in the sensitivity plot Figure 3, we took the average rate across all age groups) have a significant impact on *R*_0_, whereas the testing coverage *π* and modification factor *ι* have a minor influence on *R*_0_. The findings suggest that keeping track of the “hidden” effects of asymptomatic transmission, shortening the infectious periods, and lowering hospitalisation rates—which, in our scenario, could serve as “treatment” since we assumed that hospitalised individuals do not further contribute to infection—are nearly as crucial as decreasing contact during an outbreak. The insignificant impact of the testing rate in Figure 3 could be attributed to the negligible value of the test proportion during the initial phase of the pandemic.

Parameter *π*(*t*) represents the fraction of all infected individuals identified (tested) at time *t*. These detected individuals are assumed to isolate (or be isolated) and have reduced onwards transmission (modified by the parameter *ι* ≪ 1), whereas undetected infectious individuals (fraction 1 − *π*(*t*)) continue to spread with full transmissibility. However, improving the testing capacity is critical in reduction infection transmissibility, as shown in Figure 4.Although it may not lower the reproduction number (*R*_0_) to below one on its own, it can substantially decrease *R*_0_. However, when paired with measures like contact reduction, strict isolation, vaccination, and so on, it could potentially bring *R*_0_ below 1, thus curbing the epidemic’s spread. This is plausible because testing individuals alone without taking necessary actions towards isolation or treatment will be counter-intuitive. *Contreras et al*. [41] confirm that solely relying on testing, tracing, and isolation is inadequate for controlling the spread of COVID-19, unless these measures are integrated with additional pharmaceutical or non-pharmaceutical strategies.

Figure 8 shows how pairwise changes in key epidemiological parameters jointly shape the basic reproduction number *R*_0_. In Panel A, we vary the relative transmission rate 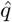 relative to the fraction of asymptomatic individuals *ρ*. The contours slope upward, indicating that an increase in either 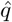 or *ρ* drives *R*_0_. A combination of both can significantly alter the trajectory of an epidemic. Panels B and C replace *ρ* with recovery rate *γ* and hospitalisation rate *δ*, respectively. If recovery and treatment is lower, and the relative transmission rate is higher, an increase transmissibility due to increased value of *R*_0_ is observed. Panels D–F repeat these explorations for the remaining parameter pairs (*ρ* vs. *γ, ρ* vs. *δ*, and *γ* vs. *δ*, respectively). In Panels D and E, increasing recovery or hospitalisation can keep *R*_0_ values low if only we can keep the number of asymptomatic individuals relatively low. This shows that when infectious individuals are removed quickly through recovery or clinical isolation, the amplifying effect of hidden (asymptomatic) transmission could be substantially blunted. Panel F illustrates that a longer duration to recovery combined with reduced hospitalization rates correlates with an increased value of *R*_0_. This emphasizes the critical role of timely clinical care. Figure 9 repeats these comparisons but now varies the scaled overall contact-mixing (*λ*) vs. each of *ρ, γ*, and *δ*. We deduce that interventions aimed at minimising contact or social interactions, in conjunction with key measures such as improving recovery through treatment, are crucial to effectively manage the spread of the disease.

By integrating age-structured contact data with a novel deprivation-decile mixing framework, this model provides a rigorous yet practical tool for understanding and controlling infectious diseases such as COVID-19. Analytical evidence and numerical findings indicate that reducing contact mixing and relative transmission rates, along with rapid recovery and treatment, is crucial for lowering the disease burden during infection outbreaks. This aligns with findings in the literature [33, 35, 40] that significantly decreasing the contact rate combined with timely enhancement of hospital treatment can lead to more effective disease control.

## Data Availability

All data produced in the present study are available upon reasonable request to the authors

## Acknowledgments

We acknowledge the support of the Institute for Global and Pandemic Planning (IGPP) at the University of Warwick. MJT is supported by the Biotechnology and Biological Sciences Research Council (BBSRC) and University of Warwick funded Midlands Integrative Biosciences Training Partnership (MIBTP) [Grant Number BB/M01116X/1].

